# Uncoupling of CSF biomarkers and clinical status in patients with a novel mutation of ATP13a2

**DOI:** 10.1101/2025.09.02.25334661

**Authors:** C Stephani, D Ewers, D Stausberg, D Rasche, A Bahiraee, B Ahmad, M Bähr, B Downie, S Hirschel, S Pauli, C Riedel, A Sachkova, G Salinas, A Schneider, C van Riesen, B Wollnik, G Yigit, KA Nave, I Zerr, S Eggert, MW Sereda

## Abstract

Cerebrospinal fluid (CSF) biomarkers are generally assumed to reflect the severity and progression of neurodegenerative disease. We report three siblings carrying a novel heterozygous ATP13A2 variant (p.Pro629Ser) who challenge this paradigm. Despite only slowly progressive cognitive decline, axonal neuropathy, and leukodystrophy on MRI, all established CSF markers of neuronal destruction were profoundly abnormal, with tau and phosphorylated tau levels exceeding even those seen in Alzheimer’s disease or Creutzfeldt– Jakob disease. Patient fibroblasts revealed lysosomal alterations and abnormal amyloid precursor protein processing, and CSF polyamine levels were strongly reduced, consistent with impaired ATP13A2 function. These cases define a new ATP13A2-associated phenotype and, most importantly, demonstrate a striking dissociation between biomarker profiles and clinical course, questioning the general validity of CSF markers as direct indicators of disease severity.

## Introduction

A range of cerebrospinal fluid (CSF) biomarkers of neurodegeneration such as types of beta-amyloid proteins, tau proteins or neurofilaments have been discovered in the last four decades and have become central to the diagnostic work-up of neurodegenerative diseases ^1,2^. Alteration of the CSF concentrations of these proteins have been thoroughly studied in the most common dementias i.e. Alzheimer’s dementia (AD), vascular dementia, and Parkinson’s disease dementia and often precede the diagnosis ^3,4^. In fact, their concentrations correlate with intracellular key pathologies (e.g. senile Amyloid plaques, neurofibrillary tangles) as well as with disease severity and prognosis ^2,5^ as demonstrated *in vivo* ^3^, and *in vitro* ^6^. Here, we present the case of three siblings in their 6^th^ and 7^th^ decade, who exhibited significant alterations of various destruction markers in the CSF along with a disturbed blood-brain-barrier. These changes are in stark contrast to a moderate and clinically rather stable phenotype. We have identified a novel variant in the *ATP13A2* gene (NM_022089.4(ATP13A2):c.1885C>T (p.Pro629Ser)) in a heterozygous state in all three affected individuals. ATP13A2 encodes a lysosomal and late endosomal P-type ATPase involved in lysosomal polyamine export. Mostly homozygous variants of ATP13A2 are linked to a range of neurodegenerative diseases ^7,8^. While we did not observe gross alterations in polyamine uptake of patient fibroblasts, reduced spermine und spermidine levels in patient CSF indicate dysfunctional polyamine kinetics.

## Case presentations

Patient 1 presented in his 50s with difficulty walking, progressive weakness of both legs that had been present for at least 6 months prior to presentation, and difficulty concentrating and articulating complex words. Neurological examination was remarkable for symmetrical mild to moderate paresis of all foot movements and a discrete choreatiform dyskinesia that had remained unnoticed by the patient and his spouse. On the advice of patient 1 (Pt. 1), his younger sibling (Pt. 2), with similar symptoms and neurological status and, thereafter another also slightly younger sibling (Pt. 3), who was clinically well and had an unremarkable neurological examination except for a very subtle dyskinesia, became patients of the department. Neuropsychological testing revealed comparable profiles in all patients with deficits in several cognitive domains, mainly reduced cognitive speed, deficits in executive function, and a reduced working and visuoconstructive memory. The MRIs showed varying degrees of leukoencephalopathy **(Figure 1)**. Nerve conduction studies were consistent with axonal motor neuropathy **(Supplement)**. CSF analysis showed a non-inflammatory status with markedly elevated total protein levels, as well as elevated levels of all markers of CSF-destruction we tested (e.g. tau, phosphorylated tau, neurofilament light, neuron-specific enolase, protein S100) and decreased beta-amyloid 1-42/40-ratios **(Figure 1, Table 1, Suppl. Figure 1, Suppl. Table 1 and 2)**. All patients showed slow but definite progress in these findings having been followed-up for up to ten years. Neither further laboratory testing, nor additional neuroimaging, nor commercially available genetic testing for genetically determined dementias provided clarifying information. Subsequently, by performing Exome Sequencing on genomic DNA obtained from all three index patients we were able to identify the heterozygous variant c.1885C>T in the *ATP13A2* gene (NM_022089.4) on chromosome 1. This variant, c.1885C>T, in ATP13A2 is predicted to lead to the substitution of a proline at the amino acid position 629 with serine (p.(Pro629Ser)). This variant was present in only 1 of > 1613548 alleles of the gnomAD database (MAF 0.000000620), in line with an autosomal dominant inheritance pattern. Sanger sequencing confirmed the presence of this variant in all three individuals **(Suppl. Figure 5)**, and *in silico* prediction of the pathogenic effect of this missense variants by different prediction tools lead to the classification as damaging (SIFT), probably damaging (PolyPhen-2), and a Combined Annotation Dependent Depletion (CADD) score of 27, indicating deleteriousness of this variant. ATP13A2 has recently been revealed to mediate ATP-driven polyamine export from lysosomes, thereby contributing to low luminal pH ^9,10^. In the experimental 3D structure of ATP13A2, the P629S substitution is located near the ATP-binding site **(Figure 1B)**, suggesting that it may impair polyamine transport function. To test for this possibility, skin fibroblasts were obtained from each patient, but showed unaltered ATP13A2 protein expression compared to human controls **(Figure 2K, Suppl.Figure 3)**. However, we identified significant decreases in expression of lysosomal markers Cathepsin B and mature Cathepsin D as well as reduced expression of full-length (FL.) amyloid-β precursor protein (APP), while the ratio of secreted Aβ/FL. APP and sAPP/FL. APP was significantly increased **(Figure 2)**. Uptake of fluorescent spermine did not differ between patient fibroblasts and those of healthy controls **(Figure 2J)**. However, CSF levels of both spermine and spermidine in one patient CSF were only 10% of the average of an internal neurological control cohort (**Suppl. Figure 2**).

**Table 1:** Cerebrospinal-fluid (CSF) Analysis.

| Group | Reference range | Patient 1 | N | Patient 2 | N | Patient 3 | N |
| --- | --- | --- | --- | --- | --- | --- | --- |
| Albumin (CSF/Serum) | <0.5 | 5 ± 1 | 6 | 2.7 ± 0.2 | 5 | 3.6 ± 0.3 | 2 |
| Tau protein | <450 pg/ml | 2397 ± 635 | 7 | 1982 ± 944 | 5 | 1807 ± 485 | 2 |
| p-Tau protein | <61 pg/ml | 404 ± 68 | 7 | 271 ± 80 | 5 | 287 ± 109 | 2 |
| Aβ-ratio | >0.5 | 0.19 ± 0.08 | 7 | 0.31 ± 0.07 | 5 | 0.25 ± 0.06 | 2 |
| NF | <540 pg/ml | 3426 | 1 | 2496 | 1 | 1958 | 1 |
| NSE | <30 ng/ml | 75 ± 5 | 4 | 58 ± 5 | 3 | 43 ± 3 | 2 |
| S-100 | <2.7 ng/ml | 7.6 ± 0.3 | 4 | 6.3 ± 1.1 | 3 | 5.3 ± 0.4 | 2 |
| Protein 14-3-3 | Negative | Positive | 2 | Borderline positive | 1 | Positive | 1 |

CSF Parameters: p-Tau = phosphorylated tau protein; Aβ -ratio = ratio between concentration of Amyloid- $\beta$ -1-42 to Amyloid- $\beta$ -1-40 multiplied with 10; NF = neurofilament; NSE = Neuron specific Enolase

**Figure 1:**
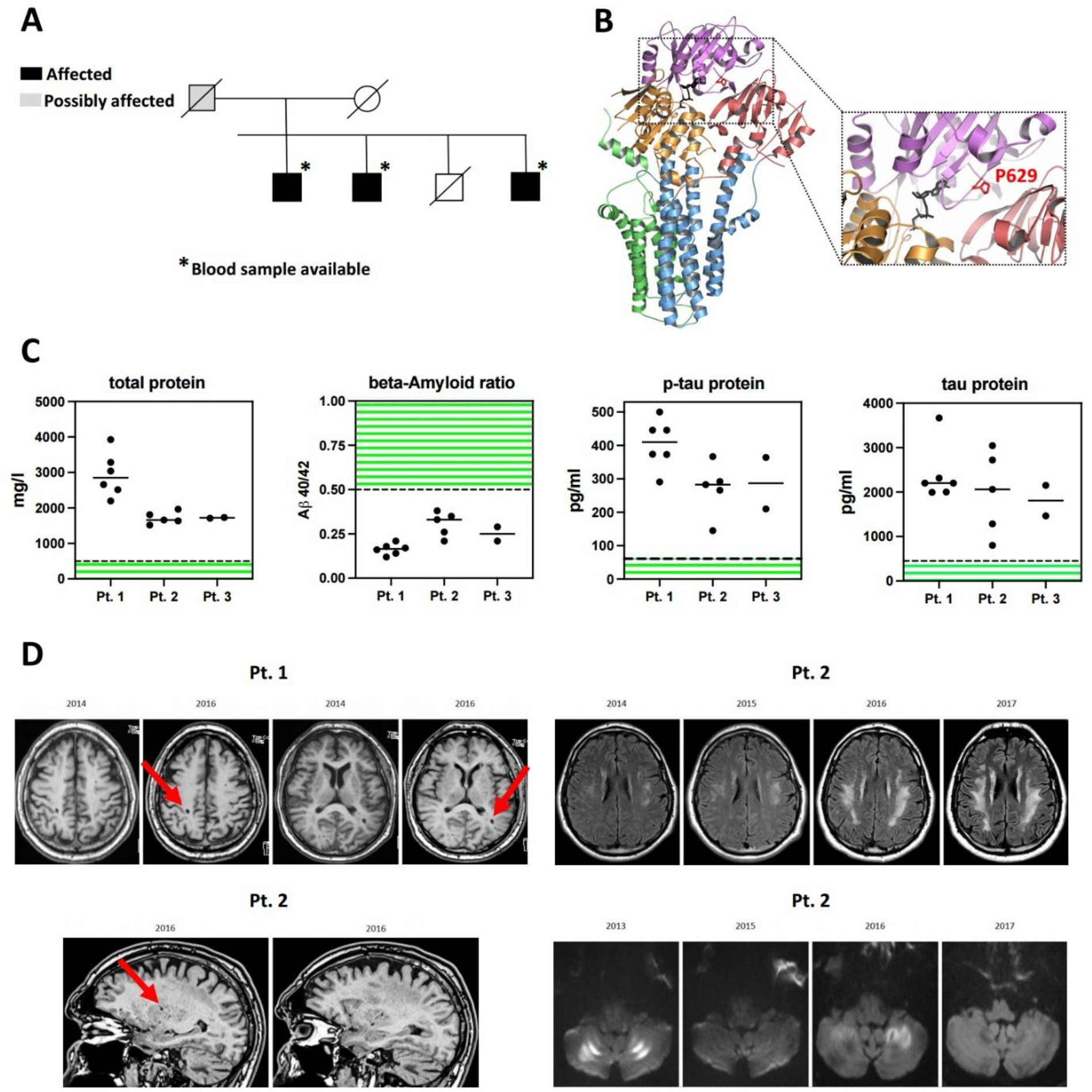
Patients carrying a heterozygous ATP13A2 mutation c.1885C>T (p.P629S). **(A)** family tree of the affected patients; the left section of the pedigree has been covered to reduce potentially identifying information **(B)** Location of mutated P629 (sticks, red) in the cryoelectron microscopy structure of ATP13A2 (PDB ID: 7VPI) within the N-domain (ribbon, light lilac) near the bound ATP analog AMPPCP (sticks, dark grey). **(C)** Repeat CSF analyses, from left to right of total protein, Aβ42/Aβ40 ratio, tau protein and phosphorylated tau protein obtained over several years; green hatches indicate the respective normal reference ranges; an accumulation of tau-values at 2000 or 2200 is due to values exceeding the upper limit of the Immunoassay-test. **(D)** Figures of two of the patients showing relevant MRI findings including small deep white matter cysts indicated by red arrows in Pt. 1 (upper left panel, T1 sequence) and Pt. 2 (lower left panel; T1 sequence) as well as progressively confluent T2 prolongation within deep white matter in Pt. 2 (upper right panel; Fluid attenuated inversion recovery imaging) (and lower right panel (C4 = diffusion weighted imaging (B value: 1000)) for selected years. P-tau = (hyper)phosphorylated tau.

**Figure 2:**
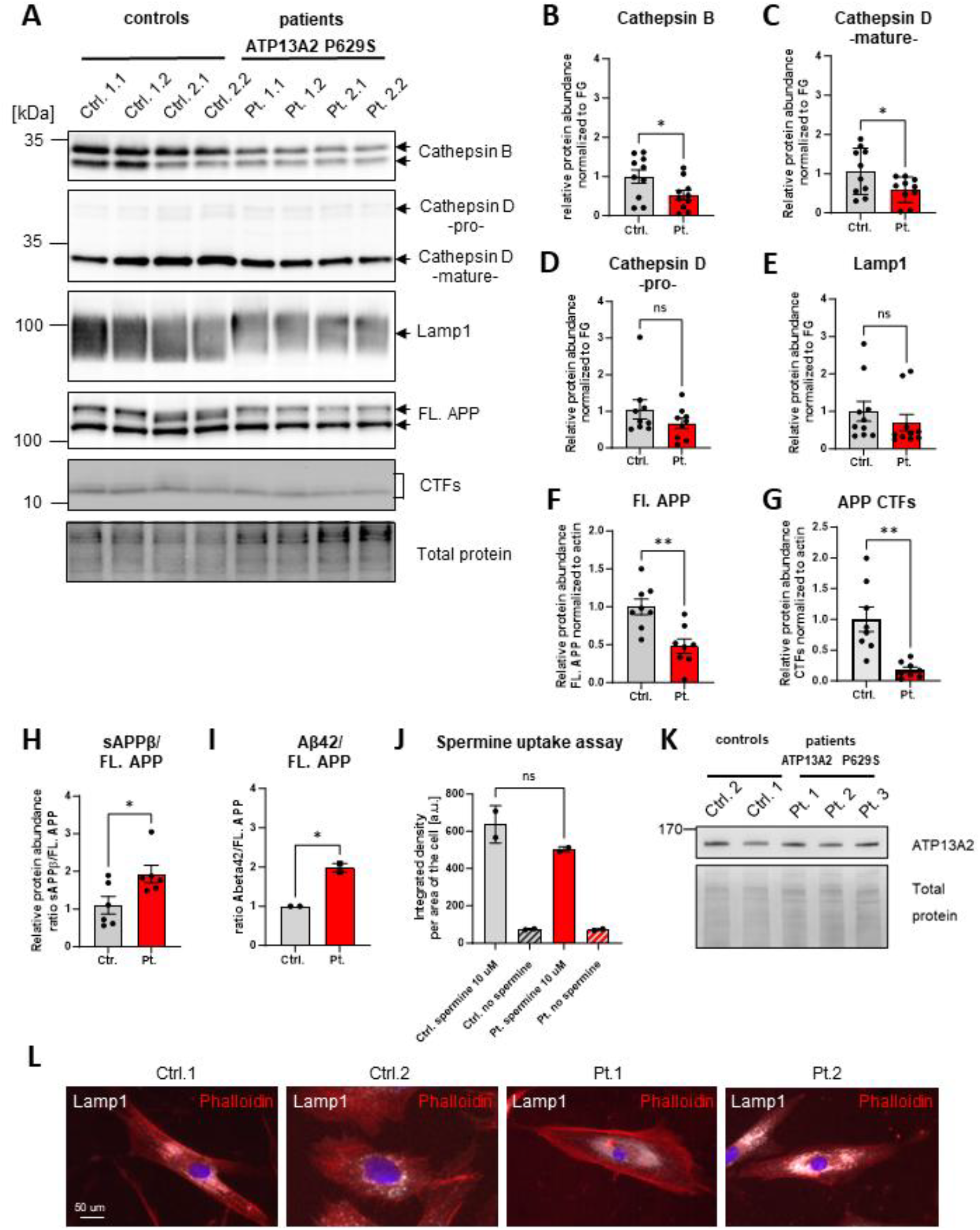
Examination of Skin fibroblasts from patients carrying a heterozygous mutation in ATP13A2: c.1885C>T; p.(P629S) versus controls. Skin fibroblasts from two slightly younger healthy volunteers and two patients were seeded and grown over night. **(A)** The cells were lysed and equal amounts of protein were analyzed via Western Blot detection. Different lysosomal proteins, Cathepsin B, Lamp1, and Cathepsin D were examined. APP and its C-terminal cleavage products were visualized with the KO validated anti-APP antibody Y188. Fast green staining enabled detection of total protein on the membrane which was used for quantification. **(B)** Cathepsin B and **(C)** mature Cathepsin D were significantly reduced in patients compared to controls while **(D)** immature Cathepsin D was not significantly altered. **(E)** Lamp1 signal was markedly reduced as a trend and a shift towards forms with a higher apparent molecular weight was visible for patients with ATP13A2 P629S protein. **(F)** Full length (FL.) APP and **(G)** its C-terminal fragments (CTF) were significantly reduced in patients versus controls while the ratio was unaffected **(Suppl. Figure 3). (H)** The ratio of secreted forms sAPPβ/FL. APP as well as **(I)** Aβ/FL. APP were significantly increased. Bars of the diagram show the mean ± SEM; N=4-6 and two technical replicates, statistics via unpaired t-Test, * p ≥ 0.05, ** p ≥ 0.01, and *** p ≥ 0.001. **(J)** Skin fibroblasts were treated with 10 µM fluorophore tagged spermine for 30 min at 37 °C and 5% CO_2_. Cells without spermine treatment served as a negative control. The cells were fixed, permeabilized and stained with primary antibody α-Lamp1 (secondary antibody goat anti-rabbit Dylight-647) as well as Phalloidin-555 to visualize actin. Nuclei were visualized using DAPI **(Suppl. Figure 4)**. Spermine uptake was decreased as a trend. **(K)** ATP13A2 protein level were not significantly altered between controls and patients in skin fibroblasts as shown via Western blot **(Suppl. Figure 3). (L)** Exemplary immunocytochemical images visualizing Lamp1, actin, and DAPI staining in human controls and patients (overlay). Bars of the diagram show the mean ± SEM; N=2, statistics via unpaired t-Test >60 cells, * p ≥ 0.05, ** p ≥ 0.01, and *** p ≥ 0.001. Scale bar is 50 µm.

## Discussion

This combination of clinical phenotype and a discrepant novel CSF status is unlike any we are aware of. All CSF parameters indicating CNS destruction were severely altered. Remarkably, in two of our index patients we measured levels of tau- as well as phosphorylated tau-protein that exceeded every single value of a large reference cohort of the CSFs of 5767 patients with dementias of various types ^3^. The values of phosphorylated tau also surpassed those of another reference cohort of more than 5000 neurological patients with various brain diseases (inflammatory, paraneoplastic, ischemic, epileptic, psychiatric, metabolic) ^11^, those seen in patients with CJD ^12^, and most of our neurochemical laboratory that also serves as German Reference Center for Prion Diseases. On the other hand, amyloid beta 1-42 and the beta-amyloid ratio 42/40 were significantly decreased, constellations typically found in patients with Alzheimer’s disease (AD), while the values of tau- and phosphorylated tau protein were much too high than those expected for patients with AD or Parkinson’s disease dementia ^13^. Likewise, neurofilament light chain ^14^, protein S100 and neuron-specific enolase (NSE) were significantly elevated in CSF **(Table 1, Supplementary Figure 1)**. Moreover, protein 14-3-3 – a CSF-marker of CJD - was elevated in all patients. Interestingly, the CSF also revealed significantly elevated total protein levels, which is uncommon in neurodegenerative diseases, where changes of CSF/serum albumin ratios are found in only 10-25% of respective cohorts and generally are mild ^15^. To conclude, the constellation of CSF findings and clinical course of our patients is incompatible with any currently known neurodegenerative disease. Patients with other pathogenic variants of ATP13A2 did either have no CSF testing or CSF that was normal “for routine parameters” ^16,17^. A neuropathological post-mortem study of a patient with Kufor-Rakeb-syndrome with a proven homozygous missense mutation of ATP13a2 did not show alpha-synuclein-, tau-, or beta-amyloid pathologies ^18^. Thus, the transferability of our CSF findings to other ATP13A2 mutations remains unclear. The symmetric MRI white matter lesions with some parietooccipital predominance **(Figure 1)**, resemble those seen in adrenoleukodystrophy, a peroxisomal disorder or metachromatic leukodystrophy, an autosomal recessive storage disorder, both of which we excluded biochemically. In addition, cystic-vacuolic lesions **(Figure 1)** found in two of our patients are a typical feature of mucopolysaccharidoses, another type of lysosomal storage disorders. But again, as opposed to cerebral and/or cerebellar atrophy leukodystrophy has not been a common finding of patients with ATP13A2-related diseases ^8,16,19,20^. In disorders of lysosomal function such as metachromatic leukodystrophy total protein and tau protein in CSF indeed may be mildly elevated ^21,22^, the latter also in patients with Niemann-Pick-disease ^23^. Hence, broadening the range of ATP13A2 related diseases, our cases may best be diagnosed as a novel type of lysosomal storage disorder.

ATP13A2 belongs to the group of P5-type ATPases (ATP13A), highly conserved transmembrane transporters present in fungi and all animals. They comprise two subgroups: P5A with one isoform only (ATP13A1) and P5B with up to four isoforms (ATP13A2-5), the number of which increased with vertebrate evolution with ATP13A2 being the oldest isoform ^24,25^. In humans, ATP13A2 is located on chromosome 1 (26 kilobases, including 29 exons), encodes a protein product with 1180 amino acids and is expressed in all tissues, but predominantly in the central nervous system ^7^. Intracellularly, ATP13A2 locates to endosomal and lysosomal membranes, promotes polyamine export into the cytosol ^10^, and lysosomal acidification via its function as H+/K+-ATPase ^9^. Currently, more than 30 homozygous or compound heterozygous mutations of ATP13A2 have been described. They are associated with a variety of neurodegenerative diseases, foremost juvenile/early-onset forms of Parkinson’s disease, especially the Kufor-Rakeb syndrome (KRS) (=PARK9) ^7^, hereditary spastic paraplegia (HSP) ^8^, and neuronal ceroid lipofuscinosis (NCL) ^26^. Symptomatic heterozygous mutations as in our cases have been reported by some e.g. in association with early manifestations of PD (e.g. 2236G>A, A746T ^20,27^), but disputed by others ^19,28^. Indeed, the variant p.P629S presented here is predicted to be located within the catalytic binding site of the ATP13A2-carrier **(Figure 2)**. Thus, this position may predispose to clinical manifestation in heterozygous states and explain the significant though non-aggressive phenotype along with an intact wildtype allele. Even though we were not able to show reduced polyamine uptake of patients’ fibroblasts as compared to fibroblasts from healthy controls this may in fact be different in cells of the central nervous system. In line with this assumption, we detected significantly reduced spermine as well as spermidine levels as measured in the CSF of one of the patients (**Supplementary Figure 2**).

## Conclusion

We present three patients with a unique clinicobiological profile consisting in a mismatch between mildly progressive clinical phenotype, leukodystrophy on MRI, alongside a novel, severe dementia-like CSF status and blood-brain-barrier dysfunction. All three patients carry a likely pathogenic new ATP13A2 mutation, a movement disorder gene coding for lysosomal polyamine and proton transporter. We propose that these cases extend the phenotype of this gene, may provide new insights into the role of lysosomal and polyamine pathways in neurodegenerative disorders and challenge assumptions about biomarkers.

## Data Availability

All data produced in the present study are available upon reasonable request to the authors

## Acknowledgment

We would like to thank Beate Veith for her excellent support regarding cell culture of the fibroblasts, Western Blot analyses, and ELISA measurements. We would like to thank medical technician Ms. Ehbrecht for expanding the first passages of the fibroblasts. We would like to thank Susan Lisa Stahmer for collecting CSF-probes for polyamine measurements. We also would like to thank doctors Anja Manig, Berenice Lammering and Jonna Meincke for participating in the patient care over the 5-years course of follow-up.

