## Supplement for "Uncoupling of CSF biomarkers and clinical status in patients with a novel mutation of ATP13a2"

* Shared first authorship

### Overview

**History and clinical examination:** A standard clinical and neurological examination was performed by one of the authors (CS) in every patient. Two of the patients have clinically and diagnostically been followed-up regularly on a near yearly basis since 2012 and 2013, respectively, and have been examined by the author on all these occasions with few exceptions. Three additional medical doctors were involved in the primary patient care over the time of follow-up so far (Acknowledgment). Neuropsychological testing: Neuropsychological testing was performed by one of the coauthors (SH) exclusively. It was done repeatedly, on about a yearly basis in two of the patients since 2012 and 2013 respectively, for 5 years and done twice in the third of the affected siblings. Neurophysiological examinations: Motor and sensory neurographies of upper and lower extremities were performed in all patients. Sensory evoked potentials of median, ulnar and tibial nerve were measured in two of the patients. Transcranial magnetic stimulation was performed in all three patients. Electroencephalography was performed in two of the three patients. Imaging modalities: We repetitively performed MRIs of the brain in all patients including T1-, T2-, FLAIR-, susceptibility-weighed-, diffusion-weighed-sequences as well as imaging of the whole spine. Additionally, 18-Fluor-deoxy-glucose positrone-emission-tomography (FDG-PET) as well as N-ω-fluoropropyl-2β-carboxymethoxy-3β-(4-iodophenyl)tropane single photon-emission computer tomography (123I-FP-CIT-SPECT) was performed in one affected patient (index patient P1). Chest X-ray was performed in two of the patients and computer-tomography (CT) of the head in one patient. Cerebrospinal-fluid analysis: Cerebrospinal fluid (CSF) was collected after taking informed consent from all patients. We collected CSF once in patient 3 and repeatedly in patients 1 and 2. Routinely CSF-serum-albumine, -IgG, -IgA, -IgM ratios were determined. Additionally, tau-protein, phosphorylated tau-protein, amyloid-beta-1-40, amyloid-beta-1-42 and their ratio were determined as well as protein S100 and neuron-specific enolase each in CSF and serum. Additionally, we looked for paraneoplastic and antineuronal antibodies in CSF. Analysis of protein 14-3-3 was amended and performed in the German reference center for Prion disease as well as the RT-quick-test for directly detecting Prion protein. Additionally, neurofilament light chains were quantified in an external neurochemical laboratory. After concluding that the most likely cause for the clinical findings in the three siblings would be a genetic alteration, we performed a series of single gene analyses of candidate genes. After these analyses were inconclusive, we performed exome sequencing on DNA of all three affected individuals, which provided some variants of interest, that we prioritized based on the cellular function of the encoded gene, and subsequently confirmed by Sanger sequencing. By a multidisciplinary evaluation process we chose a single gene mutation in the ATP13A2 gene to be the most likely candidate for pathogenicity in the three affected individuals. We then used protein Structure Prediction (alphafold®) to calculate the location of the single point variant within the protein. To perform functional studies with patient cells we obtained skin biopsies for fibroblast derivation and cultivation. These were then expanded for a series of functional assays. These focused on the analysis of APP proteolytic processing, a spermine uptake assay, and the measurement of lysosomal pH (all detailed in the Supplementary Section 2).

### Clinical evaluation per patient

**Patient 1:**

Examination: In our neurological exam we found nearly symmetrical mild to moderate paresis of foot elevation (MRC-scale 4-), more than pronation and supination (MRC-scale 4) and less so of foot flexion (MRC-scale 4+). Additionally, a mild thenar atrophy was observed but weakness of the upper extremities was formally absent. There was no evidence for disturbances of visual, auditory, olfactory or other sensory function. A very discrete possibly choreatiform dyskinesia was noted by the examiners but remained unnoticed by the patient and his spouse.

Medical history: Prior diagnoses of the patient were mild hypothyroidism that was treated with L-thyroxin, an urge-incontinence for which he was prescribed tamsulosine and sleep apnea treated with a sleep-apnea-mask.

Course: Over the course of more than 10 years gait disturbance, cognitive impairment and depression deteriorated progressively even though only slowly and mildly. At last examination the weakness of his feet-function was classified with a MRC-score of 3, the strength of the upper extremities was still unaffected and sensory examination was normal. The mild dyskinesia remained noticeable but without any significant progress. Therapeutically, the patient had received i.v.-steroids for 3 days followed by 5 days of intravenous immunoglobulins during his first stationary work-up for the differential of an inflammatory neuropathy without effect on the main symptoms. The patient in the following time had taken Modafinil due to the increased daytime sleepiness without a significant effect on this symptom. In the further course the patient was intermittently treated with pregabaline for muscle pain and occasional cramps which turned out to be very helpful. Citalopram is applied for his depression and dose has been steadily increased even though the patient doubts efficacy. Regular physiotherapy and ergotherapy were prescribed and were perceived as useful similar to two 4-week rehabilitations. In July 2025 the patient had moved into a nursing home due to progressive walking difficulties and cognitive decline.

**Patient 2:**

Examination: In our neurological examination we saw mild to moderate paresis of foot elevation (MRC-scale 4-), pronation and supination (MRC-scale 4) as well as foot flexion (MRC-scale 4). Also, thumb adduction and abduction of the little finger were mildly paretic. Correspondingly, we saw bilateral atrophy of calf and thenar. Again, there was no evidence for sensory dysfunction.

Medical history: Prior medical history was positive for arterial hypertension for which he was taking metoprolol.

Course: The patient reported a steady progress of his weakness and his physical lability including increasingly frequent falls and a change of his handwriting. Distal pareses were progressive with foot extension, -flexion, -supination and –pronation now exhibiting a degree of 1-2 on the MRC-scale. The patient became dependent on walking aids including an electric wheelchair which he is increasingly forced to take. Clinically, the mild choreatiform hyperkinesia that had been noticed at the index patient (Pt. 1) apparently had developed here as well again being unnoticed by the patient. Pharmacotherapeutically, we applied similar measures as to the elder sibling of the patient by prescribing pregabaline for muscle pain which was qualified as helpful by the patient. Similarly, as the patient had developed depression a selective-serotonine-reuptake-inhibitor (escitaloprame) was prescribed and its dose successively increased. Due to the unfavorable course the patient receives a pension for disabled persons as well. His last contact with our institution was in 2021.

**Patient 3:**

Examination: In his neurological examination there was a questionable degree of subtle dyskinesia but otherwise examination was mainly normal with normal strength, reflexes and sensory status.

Medical history: He was not taking any medication and his medical history was unremarkable.

Course: Clinically, there was no major change in neurological examination during the first years. At the last contact in July 2025, we still noticed a subtle amount of dyskinesia of which the patient was unaware. Nonetheless, a relative described increasing lack of concentration, memory deficits and personal withdrawal.

Details regarding the medical history may be accessed by contacting the corresponding author.

### Diagnostic evaluation

**Neuropsychological testing:**

Neuropsychological testing revealed deficits in several cognitive domains and a comparable profile in all patients foremost with reduced semantic, phonematic and cognitive velocity, deficits in executive function, reduced working memory as well as verbal learning and consolidation and a reduced visuoconstructive memory. Over the course of 12 (patient 1), 6 (patient 2) and 10 years (patient 3) respectively, these findings showed a mild but definite progress in all patients.

**Neuroimaging:**

MRI: We found a bilateral T2-hyperintense frontoparietal leukodystrophy/leukoencephalopathy without affection of U-fibres or basal ganglia and with decreased T2-signal in the cortex common to all three patients, most progressive and most significant in patient 2 (Figure 1) and patient 3. Additionally, there was a moderate brain atrophy in all patients (patient 1 > patient 2 > patient 3). Patient Pt. 1 also demonstrated polytopic periventricular T2-hyperintense almost cystic white matter lesions with partially significantly decreased T1-signal (Figure 1). These also developed as little cystic vacuolic looking lesions in follow-up MRIs of patient 2 (Figure 1) and patient 3. On the other hand, patient 2 demonstrated bilateral cerebellar T2 hyperintense signal alterations the presence of which apparently fluctuated over time (Figure 1). There were no signs of microbleeds in susceptibility weighed sequences at any time, no signs for vasculitis on MRA and no pathological contrast enhancement.

Computer Tomography: The cranial CT did not demonstrate significant abnormalities especially no cerebral calcifications (patient 2).

FDG-PET: The FDG-PET (patient 1) did not show any specific pattern of hypometabolism, which renders a diagnosis of Alzheimer's-disease less likely.

FP-CIT-SPECT: The FP-CIT-SPECT showed an unremarkable dopamine uptake well in line with the neurological status being unsuspicious for akinetic-rigid symptoms (patient 1).

**Electrophysiology:**

Nerve conduction studies: Neurographically, the amplitude of the compound motor action potentials (CMAP) of all motor nerves of the leg were reduced very mildly in patient 3 and remained unchanged within an interval of 10 years, more so and progressive in patient 1 and most significantly in patient 2 whose tibial and peroneal nerves have become inexcitable to neurographic stimulation over the past years. Significantly, there was no affection of the sural nerve in any patient. Likewise upper extremity nerves were not affected.

Electromyography: Electromyographically, there were acute as well as chronic signs of denervation in upper and lower extremities without affection of the back muscles in patient 1 and 2.

Transcranial magnetic stimulation: Transcranial magnetic stimulation revealed normal central nervous conduction time. Single measurements with prolonged conduction times were not reproducible on repeated examinations.

Sensory evoked potentials: Sensory evoked potentials of median ulnar, peroneal und tibial nerve were, as far as determined, normal.

EEG: The electroencephalography demonstrated a mild slowing of the posterior basic rhythm in patients 1 and 2.

**Cerebrospinal-fluid analysis:**

The CSF showed marked elevations of total protein in all patients and in repeated lumbar punctures (Supplementary table 1). Cell count was normal (<4 leucocytes /µl) in all CSF probes analyzed as were the CSF to serum ratios for IgG, IgA, and IgM. CSF-levels of destruction markers tau, p-tau, and beta-amyloids were measured with the Lumipulse® G1200 automatic Immunoassay analyzer (Fujirebio, Tokyo, Japan). All available destruction markers were pathological in these patients. Mean values and diagrams for all destruction marker determined are added in supplementary tables 2 and 3. Additional determination of neurofilament light chain was 3496 pg/ml and 2426 pg/ml respectively, while the cut-off for significant pathology of the external laboratory was <519 pg/ml. High destruction marker prompted the differential diagnosis of Creutzfeld-Jacob-disease (CJD). Indeed protein 14-3-3 was detectable and borderline detectable in patient 1 and 2, respectively. While it was negative in the first lumbar puncture in 2015 in patient 3 it became positive in the last CSF analysis in 2025. The RT-quick test as reference method to exclude CJD was negative when tested (in patients 1 and 2).

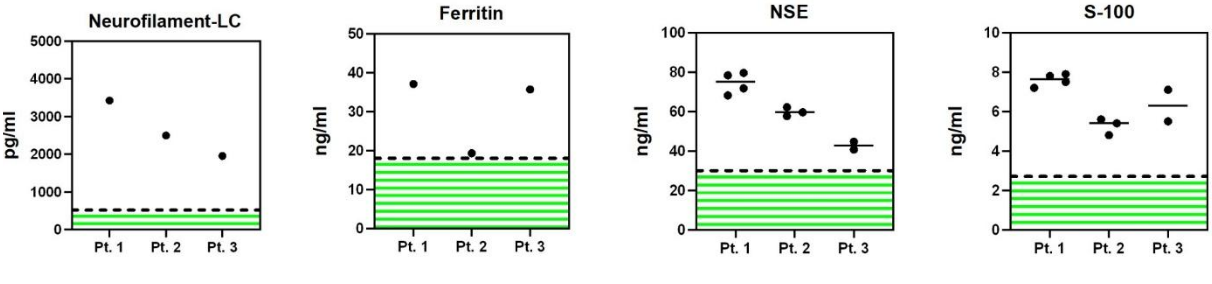

**Supplementary Figure 1:** Four figures depicting the CSF results of repeat CSF analyses, from left to right for Neurofilament light chain, Ferritin, Neuro-specific enolase (NSE), protein S-100 obtained over several years; green hatches indicate the respective normal reference ranges.

| | Year | TP in mg/l | Alb mg/l | Alb S in g/l | Alb C/S-R | L mmol/l |
|---|---|---|---|---|---|---|
| **Ref. range** | | *<500 mg/l* | *<350 mg/l* | *35-52 g/l* | | *<2 mmol/l* |
| **Patient 1** | 2014 | 2198 | 1700 | 42.7 | 3.4 | 1.6 |
| | 2015 | 2515 | 1800 | 41.9 | 4.3 | 1.8 |
| | 2016 | 3287 | 1960 | 41.8 | 4.7 | 1.6 |
| | 2017 | 2661 | 2060 | 46 | 4.5 | 1.9 |
| | 2019 | 3927 | 2990 | 67.6 | | |
| | 2021 | 3041 | 2310 | 55.9 | | |
| | 2024 | 6220* | 5790* | 40.2 | 5.6 | 3.2 |
| **Patient 2** | 2013 | 1811 | 1190 | 44.6 | 2.7 | 1.8 |
| | 2014 | 1637 | 1110 | 43.2 | 2.6 | 1.4 |
| | 2015 | 1660 | 1000 | 41.3 | 2.4 | 1.6 |
| | 2016 | 1964 | 1260 | 41.2 | 3.1 | 1.4 |
| | 2017 | 1519 | 1180 | 42.8 | 2.8 | 1.6 |
| **Patient 3** | 2015 | 1731 | 1440 | 42.8 | 3.4 | 1.9 |
| | 2025 | 1711 | 1570 | 41.4 | 3.8 | 1.9 |

**Supplementary Table 1:** CSF Results for routine CSF parameters. All routinely measured proteins (total protein, albumin, IgG, IgM, IgA) were significantly elevated in all CSF probes examined, as were CSF to serum ratios, indicating a dysfunction of the blood-CSF and/or blood-brain-barrier. CSF Lactate was normal in all patients.

| | Year | Tau (pg/ml) | p-tau (pg/ml) | Aβ 42(pg/ml) | Aβ 42/40-R | NF (pg/ml) |
|---|---|---|---|---|---|---|
| **Ref. range** | | *< 450 pg/ml* | *<61 pg/ml* | *>450 pg/ml* | *>0.5* | *<550 pg/ml* |
| **Patient 1** | 2014 | 3668 | 373 | 532 | 0.14 | - |
| | 2015 | >2000 | >500 | 442 | 0.12 | - |
| | 2016 | 2316 | 446 | 503 | 0.16 | - |
| | 2017 | 1996 | 446 | 515 | 0.18 | 3426 |
| | 2019 | >2200 | 291 | 334 | 0.17 | - |
| | 2021 | >2200 | 374 | 424 | 0.21 | - |
| | 2024 | 6220* | 5790* | 40.2 | 5.6 | - |
| **Patient 2** | 2013 | >800 | 145 | 252 | 0.21 | - |
| | 2014 | 3045 | 292 | 1613 | 0.35 | - |
| | 2015 | 1284 | 266 | 1225 | 0.26 | - |
| | 2016 | 2722 | 367 | 1291 | 0.33 | - |
| | 2017 | 2060 | 283 | 1391 | 0.38 | 2496 |
| **Patient 3** | 2015 | 1464 | 210 | 535 | 0.21 | - |
| | 2025 | 2150 | 364 | 731 | 0.29 | 1958 |

**Supplementary Table 2:** CSF Results for destruction parameters. All measurements of tau protein, the hyperphosphorylated form of tau protein (p-tau), and Neurofilament light chain were significantly elevated. Similarly, the ratio of beta-Amyloid-forms 42 to 40 (x10) was reduced in all instances, and the absolute value of beta-Amyloid 1-42 in some.

Additionally, we measured CSF concentrations of spermine and spermidine. In patient 3 we compared three measurements of different subprobes of one single CSF probe to CSF concentrations measured in a reference cohort of 34 patients with 1-3 different CSF-probes each resulting in 63 reference CSF probes. These CSF probes came from patients with lumbar or ventricular drainages due to different diseases of the central nervous system such as normal pressure hydrocephalus or subarachnoid hemorrhage. CSF concentrations of spermine and spermidine were determined by high-performance liquid chromatography coupled to tandem mass spectrometry (HPLC–MS/MS). Analyses were performed using a Shimadzu Nexera HPLC system (equipped with an SIL-30AC autosampler, LC-30AD pump, CTO-20AC column oven, and CBM-20A controller; Shimadzu, Kyoto, Japan) coupled to a Shimadzu 8050 triple quadrupole mass spectrometer (Shimadzu, Kyoto, Japan). Chromatographic separation was achieved by reversed-phase chromatography on a Brownlee SPP RP-Amide column (4.6 × 100 mm, 2.7 µm particle size), preceded by a Phenomenex C-18 guard column. The column temperature was maintained at 40 °C, and the flow rate was set to 300 µL/min. The mobile phase consisted of 0.1% (v/v) formic acid in water and an organic additive (acetonitrile:methanol, 6:1 v/v. The proportion of the organic additive was adjusted to 3%. All chromatographic and mass spectrometric parameters are summarized in Table 1. Quantification of intracellular concentrations was carried out. Concentrations were as follows:

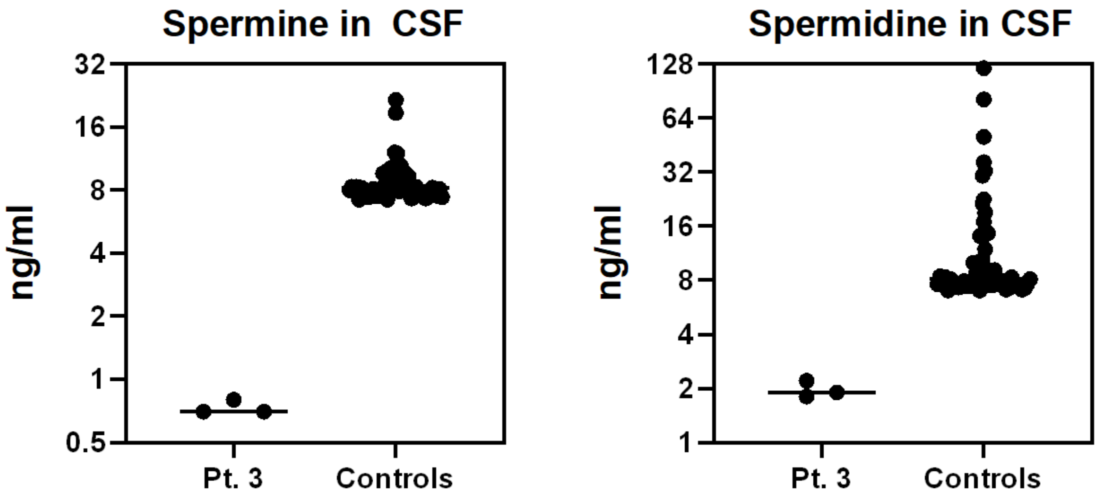

**Supplementary Figure 2:** Cerebrospinal fluid concentration of spermine (left panel) and spermidine (right panel) in patient 3 (three measurements of one CSF probe), and 34 control patients with different neurological diseases necessitating lumbar or ventricular drainages. One to three CSF samples each were obtained from these drainages resulting in 63 control measurements. Note the logarithmic y-axis.

#### Additional laboratory diagnostics:

With respect to the differential diagnosis of rare leukodystrophies we measured activities/quantities of arylsulfatase A, beta-Hexasiminidase, and very-long-chain fatty acids in patients 1 and/or 2 all of which were normal thus excluding metachromatic leukodystrophy, Tay-Sachs-disease, and X-adrenoleukodystrophy, respectively. Hu, Yo, Ri, NMDA, GM1-antibodies as well as vasculitis marker including ACE were negative as well as GM1-antibodies. No signs of sarcoidosis or tuberculosis were seen in the X-ray of the chest. Vitamin B1, B6, B12 and folic acid were in the normal range. In a prior diagnostical workup a Hashimoto encephalopathy was assumed in one patient (Patient 2) based also on a positive TPO-antibody. Nonetheless, prednisolone did not affect the state of the patient and TPO-antibody-status was normal on repeated examinations. Serology for hepatitis, Lues, and borreliosis was unremarkable in patients 1 and 2 as was blood count. HbA1c was 5.8% in patient 2 and 6.8% in patient 1.

#### Additional genetical testing:

Prior to whole-exom-sequencing we had excluded common genetic causes of motor neuropathies in one of both index patients by performing a Panel-diagnostic, and had excluded mutations in C9ORF72, Notch3 and FRAX by single gene analysis.

### Molecular biology and cell studies

**Methods:**

Skin biopsies for derivation of fibroblasts and cultivation: After obtaining informed consent from the patients we applied a round patch coated with a local anesthetic to the inner side of one upper arm. After 45-60 minutes the patch was detached, a local disinfection was performed and the skin biopsy was done by inserting a hollow needle into subcutaneous tissue and then disentangling the tissue with a scalpel. The small piece of tissue then was temporarily stored in a medium-containing flask. By this procedure we obtained fibroblasts of all three patients as well as two healthy male controls in their thirties. Tissue then was maintained in Dulbecco's modified Eagle medium (DMEM) with 10% fetal calf serum supplemented with X mM Glutamate, non-essential amino acids and streptomycin. Cultivation was done at 37°C in an atmosphere with 5% $CO^2$.

Western Blot analyses

Skin fibroblasts of patients and control patients were seeded at a number of 100,000 cells per 6 cm dish (Greiner, Cat.no. 353004) and cultured in 5 ml DMEM high glucose (Gibco) including 1% Glutamax (Gibco), 1% Penicillin/Streptomycin (Gibco), and 10% FCS. The following day, cells had a confluency of 70% and medium was changed to 2 ml volume and conditioned for 24 h. Afterwards, the media was collected. Cells were harvested and lysed in 100 µl ice cold lysis buffer [50 mM Tris/HCl, pH 7.5; 150 mM NaCl; 5 mM EDTA; 1% NP-40; 1:7 Complete Protease Inhibitor mini (with EDTA), Cat. no.: 11836153001, Roche] for 20 min on ice on a shaker. Cell debris was pelleted at 15,700 x g for 10 min at 4°C. Protein concentration was determined using the Biorad-Lowry method (Biorad Lowry, Cat. no. 5000111). For analysis of CTFs, same amounts of protein (10 µg) were analyzed on 4–20% Tris-glycine gels (Invitrogen, Cat.no. XP04200BOX). Blocking was performed in 5% milk powder in 1 x TBST. To investigate full length APP as well as its shedded fragments, cell lysates and conditioned media were separated on 8% Tris/glycine gels or 4-20% Tris/glycine gels and visualized via anti-APP antibody Y188 (1:5000, rabbit monoclonal, Abcam, Cat.no. ab32136) to detect CTFs as well as full length APP, anti-sAPPβ (1:500, rabbit polyclonal, IBL, Cat.no. 18957) to visualize sAPPβ. Secondary HRP-coupled antibodies (DAKO) were used for detection after Western blotting with ECL solution (Perkin Elmer, Western Lightning Plus ECL, Cat.no.: 0RT2655). Images were acquired via Intas Chemostar Imager at the respective Intas software and analyzed via Fiji (Image-J v1.54p) and the ratio between APP CTFs or secreted proteins and full length APP as well as protein expression levels of Cathepsin D (rabbit monoclonal, 1:1000,

Abcam), Cathepsin B (rabbit monoclonal, 1:1000, Abcam), Lamp1 (rabbit monoclonal, 1:1000, Abcam), and GM130 (mouse monoclonal, 1:1000, BD Biosciences) were quantified. Statistical analysis was performed using t-Test (Prism 10.1.2, Graphpad).

Immunocytochemistry

13 mm glass coverslips (Marienfeld, Cat. no.: 0113530) in a 24-well plate (Greiner, Cat.no. 662160) were coated with Poly-L-Lysine (Sigma) (20 µg/ml). Skin fibroblasts were seeded at a density of 10,000 cells and grown over night. The cells were treated with 1 µM BioTracker™ spermine live cell dye, which is a spermine moiety attached to a BODIPY fluorophore through click chemistry (Cat.no.: SCT250, Sigma Aldrich) for 30 min at 37 °C and 5% $CO_2$. Cells without spermine treatment served as a negative control. The cells were washed one time with 1 x PBS and fixed for 10 min at RT in 4% PFA in phosphate buffer. Subsequently, the cells were permeabilized for 7 min with 0.1% NP40. After incubation of primary antibody α-Lamp1 (Abcam) at 4 °C overnight, secondary antibody (Dylight-594, Invitrogen) as well as Phalloidin-555 (Invitrogen) were added for 1 h at room temperature (RT). Nuclei were visualized using DAPI (Roche, Cat.no.: 10 236 276 001) staining. Cells were embedded in Aqua Polymount (Cat. 18606, Polysciences) and subjected to z-stack imaging with the software Zen blue 3.3 at the microscope Axio Observer Z.1. Analysis of intensity measurements followed via Image J per area of the cell.

Analysis of APP proteolytic processing

Aβ42 in the conditioned media from skin fibroblasts of human controls and two patients has been determined via an Aβ42 ELISA from IBL (Cat.no. 27719) according to manufacturer`s instructions. 16 bit Tiff images were analyzed via ImageJ and the ratio between CTF or secreted proteins (sAPPβ and Aβ) and full length proteins was quantified. Statistical analysis was performed using t-Test.

#### Suppl. Fig 3

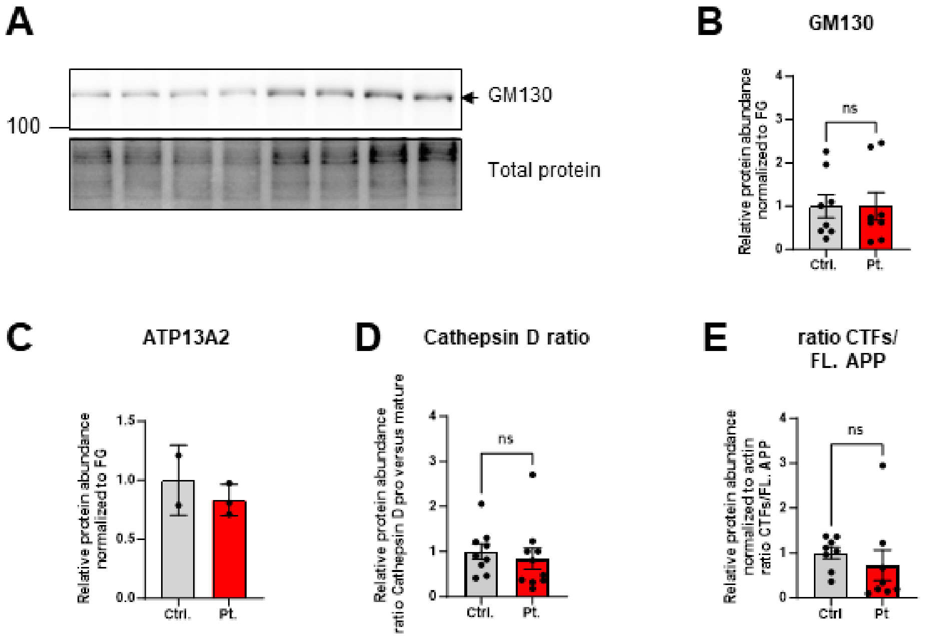

**Suppl. Figure 3: Examination of Skin fibroblasts from patients carrying a heterozygous mutation in ATP13A2: c.1885C>T; p.(P629S) versus human controls.** Skin fibroblasts from two different controls and two patients were seeded and grown over night. The cells were lysed and equal amounts of protein were analyzed via Western Blot detection. ATP13A2, GM130, Cathepsin D, and APP processing were examined (see also Figure 2). Fast green staining enabled detection of total protein on the membrane which was used for quantification. **(A)** Western Blot for cis Golgi Marker GM130 and a Fast green control is shown **(B)** quantification revealed no significant changes regarding GM130 protein expression between controls and patients **(C)** ATP13A2 protein levels were not visibly altered between patients and controls. **(D)** Cathepsin D ratio was not significantly changed in the patients **(E)** Ratio of Full length (FL.) APP and its C-terminal fragments (CTF) was not significantly altered in patients versus controls. Bars of the diagram show the mean ± SEM; N=4-6 and two technical replicates, statistics via unpaired t-Test, * $p \leq 0.05$, ** $p \leq 0.01$, and *** $p \leq 0.001$.

#### Suppl. Fig 4

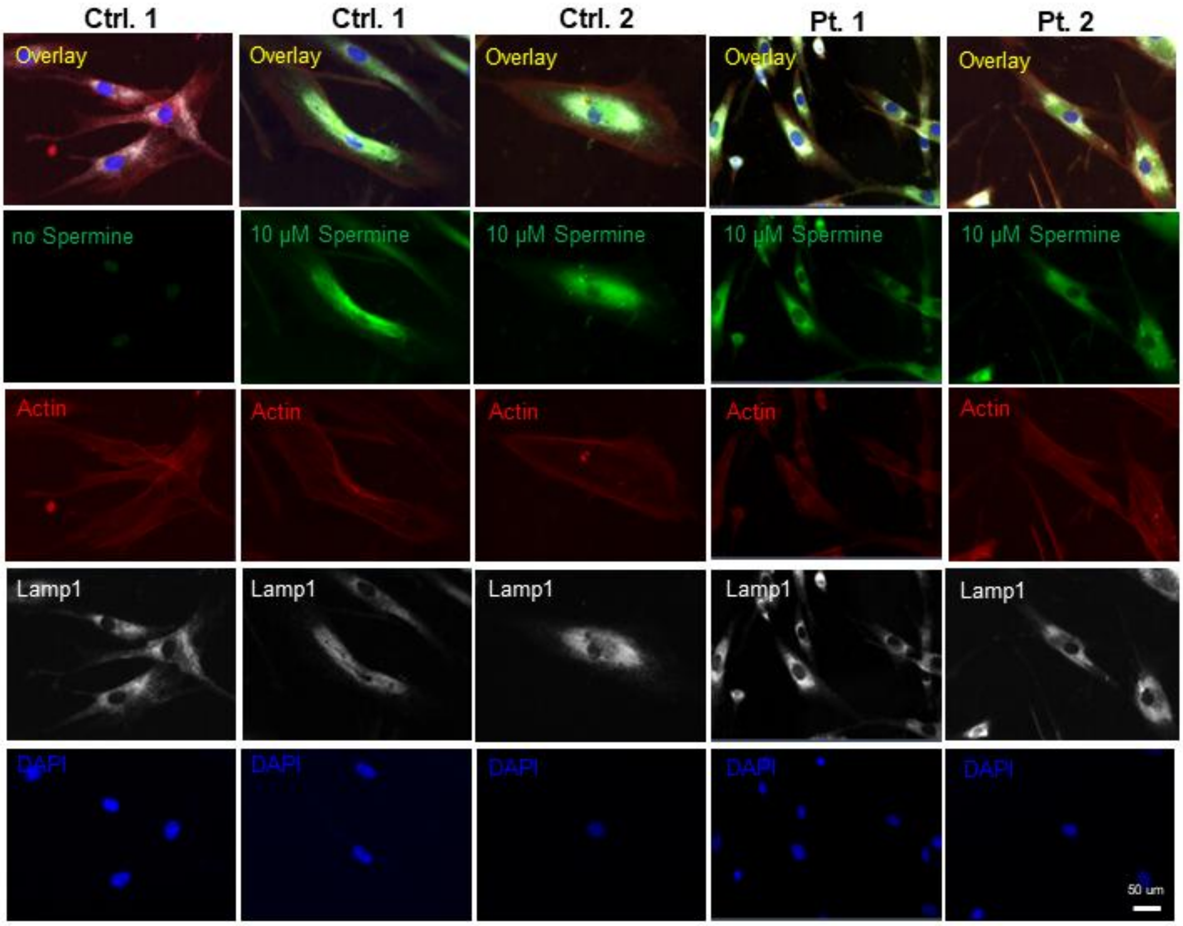

**Suppl. Figure 4: Spermine uptake assay.**

Skin fibroblasts from two different controls and two patients were seeded in 24 well plates and grown over night. The cells were treated with 10 µM fluorophore tagged spermine for 30 min at 37 °C. Cells without spermine treatment served as a negative control. The cells were fixed, permeabilized and stained with primary antibody α-Lamp1 (secondary antibody goat anti-rabbit Dylight-647) as well as Phalloidin-555 to visualize actin. Nuclei were visualized using DAPI. Exemplary images are shown. Scale bar is 50 µm.

Whole exome sequencing:

All patients gave written informed consent for the molecular genetic analyses, and DNA was extracted from peripheral blood lymphocytes by standard extraction procedures. We performed _exome sequencing (ES) on DNA of all three affected individuals using the Agilent SureSelect V6 (Agilent) enrichment kit on an Illumina HiSeq4000.[29,30] ES data analysis and filtering of mapped target sequences was performed using the 'Varbank' exome analysis pipeline of the Cologne Center for Genomics (CCG, University of Cologne, Germany) and we obtained a 20fold coverage in >90% of target sequences. We analyzed the ES data for variants with a coverage of more than 6 reads, an allele frequency of 20 - 80% in all three affected individuals, a minor allele frequency (MAF) <0,5% in the gnomAD database (v4.1.0), a predicted impact on protein function (including nonsense, splice-site, coding indel, and missense variants), and consistency with a recessive or dominant (heterozygous variants with a MAF <0.001%) mode of inheritance. Using this strategy, we were able to identify the heterozygous variant c.1885C>T in the *ATP13A2* gene on chromosome 1. This variant, c.1885C>T, in ATP13A2 is predicted to lead to the substitution of an proline at the amino acid position 629 with serine (p.(Pro483Ser)). This variant was present in only 1 of > 1613548 alleles of the gnomAD database (MAF 0.000000620), in line with an autosomal dominant inheritance pattern. Subsequent Sanger sequencing confirmed the presence of this variant in all three individuals, and *in silico* prediction of the pathogenic effect of this mis-sense variants by different prediction tools leads to the classification as damaging (SIFT), probably damaging (PolyPhen-2), and a Combined Annotation Dependent Depletion (CADD) score of 27, indicating deleteriousness of this variant.
Remaining variants were classified using different *in silico* prediction tools, prioritized based on the cellular function of the encoded gene, and subsequently confirmed by Sanger sequencing.

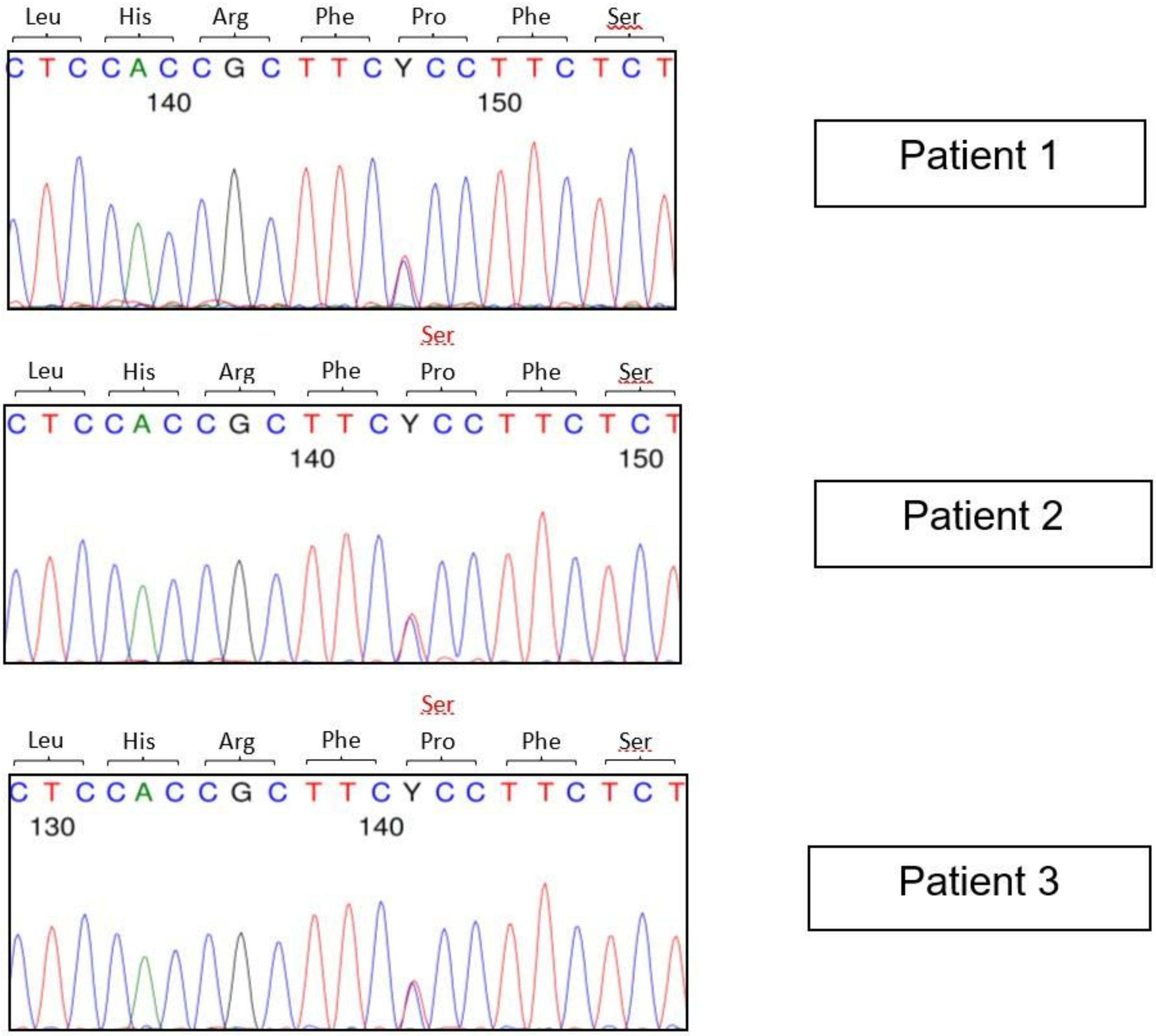

**Supplementary Figure 5:** Sanger-sequence analysis of the mutated region demonstrating a single point mutation at position 1885 resulting in an amino-acid change from proline to serine at codon 629 (c1885C>T; P629S).
